# Hidden Scars: Prevalence of adverse childhood experiences (ACEs) among medical students at the University of Khartoum, Sudan

**DOI:** 10.64898/2026.01.21.26344586

**Authors:** Somia Tawfik, Alaa T. Omer, Kamil Mirghani Shaaban

## Abstract

**Background:** Adverse Childhood Experiences (ACEs) are widespread and have detrimental long-term effects in many aspects including physical and mental health, education and employment. Medical students are presumed to have had high rates of exposure and suffer consequences that might hinder their progression in undergraduate or postgraduate level.

**Objective:** The study aimed to determine the prevalence of adverse childhood experiences among different sociodemographic groups of medical students and elicit outcomes in self-perceived physical and mental health.

**Methods:** This was a descriptive cross sectional study, carried out among 326 medical students selected by stratified and systematic random sampling, using online self-administered questionnaire (ACE-IQ), The data was analyzed using statistical package for social sciences (SPSS), Pearson chi-square test was performed to measure associations and binary logistic regression to determine the significance and odds that a student exposed to childhood trauma or ACEs is likely to report negative effects on their physical health as well as mental health.

**Results:** 81.9% of students reported at least one ACE and 23.9% reported > 4. Male gender, age group 18-21, medical students raised in a rural setting, and low family income sociodemographic groups demonstrated higher prevalence. The study noted a significant association between rising ACE scores and expressing negative effects on mental health (OR: 1.359, p value: .000, 95% CI:1.195-1.546) and Physical health (OR: 1.177, p value: 0.01, 95% CI: 1.040-1.333).

**Conclusions:** ACEs are highly prevalent among medical students, with 4 out of every 5 students being exposed and only one being unexposed. In addition, increasing exposure to adversity was associated with negative effects on physical and mental health in a graded exposure-effect manner.

## 1. Introduction

According to the Centers for disease control and prevention (CDC), Adverse Childhood Experiences (ACEs) interchangeably termed Childhood Trauma are a group of unfavorable potentially traumatic experiences taking place in an individual’s childhood from 0 to 17 years of age(1).

Childhood is pivotal for development and determinant of adult health. Positive experiences help reinforce developing bio systems. Conversely, adversity can increase morbidity and mortality(2). The resultant child’s health is comprised of the interaction between biological and environmental factors(1).

Studies elucidate increased risk for common physical diseases in poly-victimized individuals (exposed to 4 or more ACEs), this indicates that the more a child is exposed to ACEs the worse the outcome is, a concept referred to as a dose-response relationship(3). This has also been established between ACEs, impaired mental health and social difficulties ln adulthood(4).

Current literature presumes that mental health struggles are more common among medical students than other students and the general population(5). One established causative factor for mental health difficulties in general population is history of exposure to childhood trauma. It is speculated that students with history of ACEs prefer to enroll in medical schools due to the outlook that caring for others is therapeutic for themselves(6).

The burden of ACEs is large and global, extensive literature shows childhood trauma exposure is widespread across populations and a substantial minority of children experience multiple ACEs. It is estimated that one billion children worldwide-over 50% of all children have been exposed to some sort of violence in 2015(7). Furthermore, the occurrence of the COVID-19 pandemic and implementation of the lockdown and home quarantine measures is expected to increase children’s exposure to ACEs both at home and online(8).

ACEs affect the individual’s productivity and in turn influence earnings per capita and the economy of the country(9).

A study conducted in the USA found that The Quality Adjusted Life Expectancy (QALE) decreased by 9.5 years or 17% in individuals reporting more than 3 ACEs(10).

The medical field is quite demanding, despite this, medical students are often seen as privileged and acquire natural capacity to withstand challenges and stressors that affect the public. However, significant rates of depression, burnout, and suicidal ideation among trainees indicate an underlying vulnerability that precedes their medical education(11).

Growing research suggests Adverse Childhood Experiences (ACEs) as a reason for this that is often overlooked. The connection between cumulative childhood trauma and poor adult health is well-established in the general population, nonetheless, the prevalence of ACEs within medical students remains under-investigated. Recognizing this knowledge gap is crucial, as unresolved childhood adversity can have a negative impact on future doctors’ wellbeing, professional identity, and ability to care for patients.

This study aimed to estimate the total prevalence of ACEs among medical students in University of Khartoum, Sudan during the first 18 years of life, In addition, it was set to determine the prevalence of each ACE category separately, to compare prevalence among certain demographic groups and to assess current self-perceived physical and mental health as a subjective measure of impact/ outcome.

## 2. Methodology

### 2.1 Study design

This study was descriptive cross-sectional, institutional based among medical students.

### 2.2 Study Setting

Faculty of Medicine, University of Khartoum, Khartoum, Sudan.

It is the oldest and largest University in Sudan, it was founded in 1902 as Gordon Memorial College, later renamed in 1956 after the country gained its independence(12).

Students: represent high achieving students in high school certificate exam who have been accepted via degrees’ competition, largely diverse group representing different states in the country. Number of Students enrolled in Faculty of Medicine at the time of the study was 2341.

### 2.3 Study Population

Medical Students from 1^st^ to 6^th^ grade.

^*^Inclusion Criteria:

1. Students who are currently enrolled in Faculty of Medicine at the time the study was conducted.
2. Students aged 18 and above.

Sampling

### 2.4 Sample Size

The sample size was calculated using the equation for known population size (Slovin’s formula): **n = N/ (1+Ne 2)** with assumptions of:

N: population size = 2341

e: margin of error = 0.05, Confidence Level 95%

Sample size (n) = 2341/ 2341+1(0.05^*^0.05) = 341.6 =app. **342**

### 2.5 Sampling Method / Technique

To guarantee representative participation from the whole student population, the study used a proportionate stratified random sampling technique. Based on academic grades, the population was split into seven different strata (Years 1 through 6). The first-year cohort was then split into two different entrance batches (Batch 96 and Batch 97). Each subsample’s size was determined to be proportionate to the weight of its respective stratum within the overall population (N = 2341).

A methodical sampling strategy was applied within each stratum. Based on the ratio of the entire population to the intended sample size (n = 342), a sampling interval (K) of 6 was determined using a complete sampling frame of all registered medical students. Every sixth student was chosen after the first subject for each stratum using a random integer generator (Random.org) (13).With individual strata contributions ranging from 46 to 52 people, this procedure produced a final cohort of 342 students, precisely mirroring the 13.3% to 15.3% population distribution of each academic year.

### 2.6 Data Collection

Instrument: Data was collected using Self-Administered Questionnaire, containing close-ended questions. After explaining the questionnaire and taking informed consent from each participant. The Questionnaire used is the Adverse Childhood Experiences International Questionnaire (ACE-IQ)(14) with a few edits to suit medical student participants and the age group. The section about marriage and intimate partner violence was deleted and 2 questions about self-perceived health were added.

Questions cover family dysfunction; physical, emotional and sexual abuse and neglect by parents or caregivers; peer violence; witnessing community violence, and exposure to collective violence(14). It contains 43 items asking about 13 categories, the answers are then dichotomized, and the individual is either given a value of 1 to indicate exposure to an ACE category or 0 to indicate non-exposure and the individual’s final ACE score ranges from 0 to 13. Higher scores indicate exposure to multiple ACE categories.

Information collected included: Demographic info (gender, age, growth environment and family income), questions on relationship with parents, family environment, adverse experiences the participant went through during the first 18 years of life, peer violence, physical fights, witnessing community violence and exposure to war. Besides added questions about self-perceived physical and mental health.

Online Google forms with Questionnaire and consent were created and the link was sent to selected students directly from the Author through social media platforms. Data collection took place between Sunday, 24th January 2021 and continued for 3 weeks, ending on 14th of February. Number of responses gathered within data collection time frame was 326 (95.3% response rate).

### 2.7 Data Management & Analysis

Collected data was coded using a code book provided for the ACE-IQ for analysis using Statistical Package for Social Science (SPSS) ® version No. 23 software for statistical analysis(15). Simple descriptive statistics: Frequencies, percentages and chi squire test & Fisher exact probability test were used. And binary logistic regression was used to determine the significance of perceived physical and mental health outcome.

### 2.8 Ethical Consideration

The study was conducted in line with the ethical principles for medical research involving human subjects as outlined in the Declaration of Helsinki(16). Prior to data collection, the study protocol was reviewed and approved by the research ethics committee in the department of Community Medicine, University of Khartoum, Ethical Approval number: [COMMED 2021-92-31]. All participants were provided with information explaining the study’s objectives, the voluntary nature of participation, and their right to withdraw at any stage. Informed consent was obtained from all subjects before their inclusion in the study. Data were anonymized to ensure privacy and confidentiality, and no personally identifiable information was gathered or kept in the final dataset.

### 2.9 Funding

This research did not receive any specific grant from funding agencies in the public, commercial, or non-profit sectors.

## 3. Results

### 3.1 Socio-demographic characteristics of medical students participating in the study

The study included 326 participants, of which 224 were females (68.7%). The overall mean age was 21.47 years, with a standard deviation of 2.174 (Table 1).

**Table 1.** Summary of sociodemographic characteristics of participating medical students (N=326).

| Characteristic | n | (%) |
| --- | --- | --- |
| **Gender** |  |  |
| Male | 102 | 31.3% |
| Female | 224 | 68.7% |
| **Age in years, mean(SD)** | 21.47(2.174) |  |
| 18-21 | 176 | 51.2% |
| 22-25 | 152 | 46.6% |
| More than 25 | 7 | 2.1% |
| **Batch Number** |  |  |
| 91 | 49 | 15% |
| 92 | 49 | 15% |
| 93 | 46 | 14.1% |
| 94 | 49 | 15% |
| 95 | 43 | 13.2% |
| 96 | 43 | 13.2% |
| 97 | 47 | 14.4% |
| **Growth Environment** |  |  |
| Urban | 286 | 87.7% |
| Rural | 40 | 12.3% |
| **Family Income** |  |  |
| High | 50 | (15.3% |
| Average | 258 | 79.1% |
| Low | 18 | 5.5% |

### 3.2 Adverse Childhood Experiences prevalence among medical students in University of Khartoum

#### 3.2.1 Overall Prevalence estimate

ACE-IQ data were processed using a frequency-based analysis. Participants were categorized as ‘exposed’ (1) or ‘non-exposed’ (0) for each trauma category based on whether their reported frequency of exposure met the defined threshold for that specific ACE dimension. Then all of the 13 categories will sum up to the individual’s ACE score ranging from 0 to 13. The overall prevalence represents individuals who were exposed to at least one ACE category (ACE score > 0). 81.9% of medical students were exposed to at least one ACE category, while only 18.1% were not exposed to any of the 13 categories.

- ACE Score groups and their frequency

The majority of medical students were exposed to a few ACE categories ranging from 1 to 3, While a significant proportion was poly-victimized under the effect of 4 or more categories (Figure 1).

**Figure 1.**
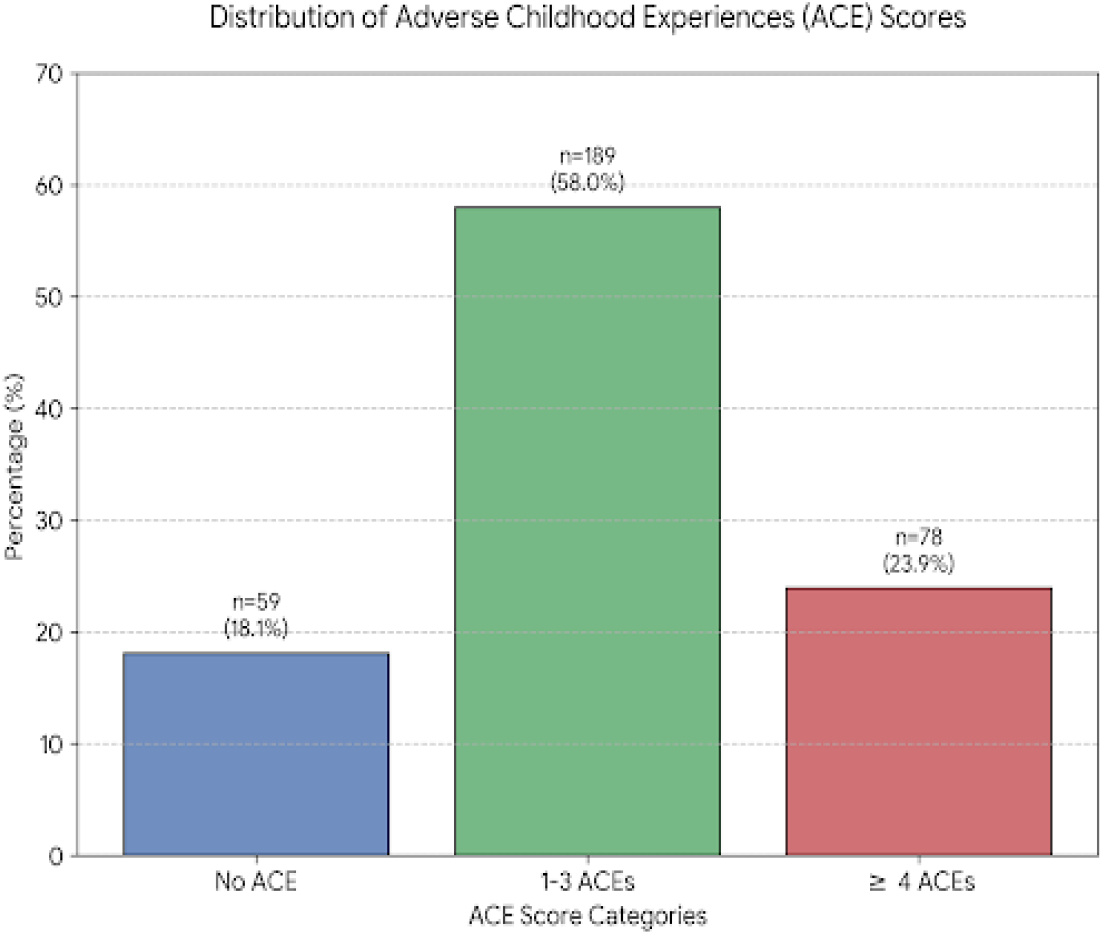
percent of ACE scores groups of medical students in University of Khartoum (N=326).

#### 3.2.2 Prevalence of ACE Categories

Overall, the most prevalent ACE category among medical students was household member treated violently, while the least prevalent was physical neglect (Table 2).

**Table 2.** Prevalence estimates for all 13 ACE categories both in count and percent among medical students in Khartoum University (N=326).

| ACE Category |  | Count | Percent |
| --- | --- | --- | --- |
| Physical abuse | Exposed | 46 | **14.1%** |
|  | Not Exposed | 280 | 85.9% |
| Emotional abuse | Exposed | 64 | **19.6%** |
|  | Not Exposed | 262 | 80.4% |
| Contact sexual abuse | Exposed | 87 | **26.7%** |
|  | Not Exposed | 239 | 73.3% |
| Alcohol and/or drug abuser in the household | Exposed | 12 | **3.7%** |
|  | Not Exposed | 314 | 96.3% |
| Incarcerated household member | Exposed | 24 | **7.4%** |
|  | Not Exposed | 302 | 92.6% |
| Someone chronically depressed, mentally ill, institutionalized or suicidal | Exposed | 25 | **7.7%** |
|  | Not Exposed | 301 | 92.3% |
| Household member treated violently | Exposed | 143 | **43.9%** |
|  | Not Exposed | 183 | 56.1% |
| One or no parents, parental separation or divorce | Exposed | 66 | **20.2%** |
|  | Not Exposed | 260 | 79.8% |
| Emotional Neglect | Exposed | 103 | **31.6%** |
|  | Not Exposed | 223 | 68.4% |
| Physical Neglect | Exposed | 9 | **2.8%** |
|  | Not Exposed | 317 | 97.2% |
| Bullying | Exposed | 32 | **9.8%** |
|  | Not Exposed | 294 | 90.2% |
| Community Violence | Exposed | 99 | **30.4%** |
|  | Not Exposed | 227 | 69.6% |
| Collective Violence | Exposed | 85 | **26.1%** |
|  | Not Exposed | 241 | 73.9% |
|  | Total | 326 | 100% |

#### 3.2.3 ACEs among different demographic groups

Using cross tabulation and Chi-square test, a statistically significant gender difference was found in physical abuse, Community Violence and Collective Violence. In all 3 **males** had higher prevalence (Table 3).

**Table 3.** Association between ACE categories and Gender, Age, Growth Environment and Family Income among medical students (N=326).

|  | Pearson Chi-Square test |  |  |  |
| --- | --- | --- | --- | --- |
|  | *Gender | *Age | *Growth<br>Environment | *Family<br>Income |
| Physical abuse | .023 | .009 | .104 | .176 |
| Emotional abuse | .552 | .257 | .950 | .544 |
| Contact sexual abuse | .631 | .158 | .375 | .085 |
| Alcohol and/or drug abuser in the household | .055 | .236 | 1.000 | .145 |
| Incarcerated household member | .816 | .233 | .515 | .777 |
| Someone chronically depressed, mentally ill, institutionalized or suicidal | .413 | .281 | 1.000 | .183 |
| Household member treated violently | .305 | .193 | .599 | .043 |
| One or no parents, parental separation or divorce | .624 | .788 | .039 | .066 |
| Emotional Neglect | .953 | .540 | .145 | .467 |
| Physical Neglect | .062 | .441 | .305 | 1.000 |
| Bullying | .684 | .000 | 1.000 | .019 |
| Community Violence | .019 | .037 | .496 | .000 |
| Collective Violence | .000 | .244 | .339 | .028 |

Physical abuse, bullying and community violence showed statistically significant differences. In all 3 categories, the **age group 18-21** had higher prevalence.

One or no parents, parental separation or divorce showed statistically significant difference. Individuals who were raised in **rural** settings had higher prevalence of parental separation.

Four ACE categories were statistically significant when compared to different family income groups: Household member treated violently, Bullying, Community Violence and Collective Violence. In all 4, students who were raised in **low-income** families had higher prevalence.

#### 3.2.4 Effects of exposure to ACEs on Self-perceived physical and mental health

Adverse childhood experiences were found to have statistically significant negative impact on both; a point increase in ACE score lead to app. 1.2 times increase in likelihood of reporting negative physical impact, and 1.4 times increase in likelihood of reporting negative mental impact (Table 4).

**Table 4.** The impact of adverse childhood experiences (ACEs) upon self-perceived physical and mental health among medical students in Khartoum University 2021 (N=326).

| impact | Odd ratio | P value | 95% C.I for odd ratio |  |
| --- | --- | --- | --- | --- |
|  |  |  | Lower | upper |
| Negative Physical impact | 1.177 | 0.01 | 1.040 | 1.333 |
| Negative Mental impact | 1.359 | .000 | 1.195 | 1.546 |

## 4. Discussion

This is the first study to provide a prevalence estimate addressing Adverse Childhood Experiences comprehensively; including all 13 categories in the ACE-IQ in Sudan. Analysis of the ACE-IQ using the frequency version showed that overall, 81.9% of medical students in Khartoum university, Sudan 2021, reported at least 1 ACE. This is relatively higher than estimates reported by developed countries. For example, in a study conducted among 86 third year medical students from a medical school on the west coast of USA, 51% reported at least one ACE(11). In addition, in a German study including 1466 university students, 73.8% was the overall ACE prevalence estimate(17).

Conversely, a study from Vietnam, a developing country revealed that 76 % had been exposed to at least 1 ACE(18). Similar results were found by a recent study in Saudi Arabia including 931 adults in Riyadh, of them 82% showed exposure to at least one ACE(19). Even higher estimates were reported by a study in Malawi among adolescents where less than 1% only reported no ACE, that being approximately 99% prevalence(20).

The results of this study are rather on the higher side, reasons for this might be the difference in socioeconomic circumstances in comparison with the developed countries where individuals and families suffer from hardships and unsettlements that contribute to adversity, for example the period of social unrest in Sudan starting 2018 might have contributed to witnessed community and collective violence thus raised ACE scores.

Another reason might be different measuring instruments and analysis methods, for example the US medical student study measured only 10 categories, whereas this study measured 13. Also, recall bias might have played a role, as the ACE-IQ is retrospective and focuses on the outcomes of adversity, the minimum age of participants required is 18 years old, 98% of participants in this study aged between 18-25 years thus might have better recall of past events than older populations.

The most prevalent ACE categories in this study were: Household member treated violently 43.9%, Emotional Neglect 31.6%, Contact sexual abuse 26.7%, community and collective violence 30.4% and 26.1%. The most prevalent categories differed from country to country though some were shared like community violence and household member treated violently. It must be noted that the prevalence of sexual abuse being 26.7% (A quarter of all participants) in this study was particularly higher than in literature, compared to the original Kaiser Permanente ACE study where sexual abuse prevalence was 22%(2), The German 12.3%(17), Another US study conducted in 23 states study found it to be 11.6%(21), The Malawian study 6.52%(20). A possible reason for this is the threshold for defining someone as “sexually abused” in the ACE-IQ frequency version of analysis, where any affirmative answer including ‘once’ to any of 4 questions is enough to dictate sexual abuse, asking about whether or not someone touched/ fondled them, made them touch their body in a sexual way, attempt oral/anal or vaginal intercourse as well as forcing to have sexual intercourse against an individual’s will(14).

Sociodemographic distribution of ACEs varied between different studies. This study showed that males had higher prevalence than females, 84.3% vs 80.8%, this is consistent with Saudi Arabian study(19), and contrary to the US medical students’ study where females had higher prevalence(11). This might be due to cultural differences that are tougher on males or less willingness of females to disclose traumatic experiences.

Regarding Age 18-21 years age group had the highest prevalence, this is somewhat similar to a Serbian study among 2,792 nationally representative sample of adults aged 18–65 years old, where younger participants (age 18 to 29 years) reported higher exposure(22), furthermore, agreeing with the results of the original Kaiser-CDC ACE study that showed a markedly lower prevalence among participants who were aged 45 and older compared to other age groups(2). However, this could again be due to stronger recall in younger individuals than older ones, in addition more recent estimates are needed taking into account both older and younger age groups to properly make that comparison.

Regarding growth environment, students who were raised in a rural setting had slightly higher prevalence than those who were raised in urban setting (82.5% vs 81.8%) this is supported by the Malawian study that was conducted in a rural setting(20), and opposed by the Serbian study(22). One possible reason for higher ACEs in rural settings in this study would be the financial hardships faced as less resources are there and relatively more stressful environments due to unavailability of services.

Low family income was associated with higher prevalence of ACEs, this was in line with the 23 United States study among 214,157 adults using the behavioral risk factor surveillance system data, where low income (<15 000$ per year) was associated with higher ACEs(21). The German study also found that a high family socioeconomic status was a consistent protective factor against most ACEs(17).

This study found strong associations between exposure to ACEs and negative self-perceived health, both physical and mental, but mental more so than physical. Binary logistic regression values were as follows: Physical (OR: 1.177, p value: 0.01, 95% CI: 1.040-1.333) mental (OR: 1.359, p value:.000, 95% CI:1.195-1.546). This agrees with the US third year medical school study where all of the students who experienced 4 or more ACEs reported moderate or significant effect on their mental health (P = <.0001)(11), however the association for physical health was not statistically significant(11). These associations have also been proven using objective measures in many studies including the original ACE study(2) the Malawian study(20), and the Saudi Arabian study(19).

## 5. Conclusions

The study concluded that approximately 8 out 10 medical students in University of Khartoum, Sudan have been exposed to at least one category of adverse childhood experiences before the age of 18. The top reported ACE categories were: household member treated violently, Emotional Neglect, Community Violence, Contact Sexual Abuse and Collective Violence. ACEs were common across all sociodemographic groups, but some groups suffered particularly higher rates than others, those include: Males, 18-21 years’ age group, those who were raised in a rural setting and those who had low family income.

The more ACEs an individual was exposed to, the more likely they were to report impaired mental and physical health (dose-response relationship).

## Data Availability

Excel data sheet link will be shared after the acceptance of the manuscript

## 6. Limitations of the study

Due to limited time and resources and the situation with COVID-19 pandemic, this study did not include the general population but instead, confined to medical students in Khartoum University, Sudan. Nonetheless, students in faculty of medicine, University of Khartoum come from diverse backgrounds, so they represent different components of the general population.

Methodological limitations of the study being retrospective, so recall bias might affect results, and the cross-sectional nature does not allow for establishment of causality between ACEs and outcomes.

## 7. Recommendations

To mitigate the negative effects of ACEs coordinated efforts need to be established on multiple levels in society. Internationally, knowledge, experience and feasible practices to be shared in a global network. Nationally, ACEs must be recognized as a public health issue and appropriate measures need to be taken in primary prevention, as ACEs outcomes are resource consuming and difficult to treat. On Faculty level, it is crucial to provide medical students – the future healthcare providers—with psychological support and safe spaces to process past adversities, particularly the community violence experienced during the 2018–2019 social unrest. Finally, emphasis must be placed on parental counselling, to break the cycle as ACEs are transgenerational and the offended tend to offend.

## 8. Acknowledgements

I would like to extend my sincere gratitude to the department of Community Medicine, University of Khartoum for allowing me the opportunity to conduct this study and guiding me through the process. In addition, I wish to honor the memory of Prof. Asmaa Abdulaal her support was invaluable, may she rest in peace.

Special thanks to my friends and support system, Farida Ahmed, Soumya Abdulaal and Sondos Ahmed, without the help of this work would not have been completed. My colleagues who generously offered me their time and assistance in all phases of this study, whether they participated and filled out the questionnaire, supported me with encouraging words, or helped me navigate the experience.

## 9. Author’s Contributions

Somia Tawfik is the first and corresponding author and contributed to study conceptualization, data curation, formal analysis, investigation, methodology, project administration, writing the original draft as well as reviewing and editing.

The following authors have also contributed to the study via different roles: Alaa T. Omer, Kamil Mirghani Ali Shaaban. They each performed data curation, formal analysis, investigation, supervision, writing-original draft and writing-reviewing and editing.

